# Persistent intestinal abnormalities and symptoms in cystic fibrosis: The underpinning mechanisms impacting gut health and motility. Protocol for a systematic review

**DOI:** 10.1101/2020.07.13.20144808

**Authors:** R.J. Marsh, C. Ng, G. Major, D.W. Rivett, A. Smyth, C.J. van der Gast

## Abstract

**Background:** Patients with cystic fibrosis (CF) are characterised by abnormalities of the intestinal tract relating to gut motility and physiological issues, with daily symptoms of disease including abdominal pain, flatulence, bloating, and constipation. With improvements in respiratory outcomes, a shift in disease manifestations has highlighted the prevalence of the gastrointestinal-related problems associated with CF, yet most therapies currently in clinical use for the gut symptoms of CF have been repurposed from other disease indications and have not been developed with a knowledge of the mechanisms underpinning gastrointestinal disease in CF. Increased attention towards the role of intestinal inflammation and microbial dysbiosis in the CF population warrants a comprehensive knowledge of these aspects alongside the increased luminal fat content, dysmotility, and small intestinal bacterial overgrowth (SIBO) resultant of the primary consequences of CFTR dysfunction (disrupted fluid secretion and pancreatic insufficiency), and how they contribute towards the intestinal complications of CF disease.

**Methods and Study Design:** We will conduct a systematic review to comprehensively address our current understanding of the primary consequences of CFTR dysfunction, and their subsequent secondary effects that contribute towards the disruption of gut motility, health, and associated symptoms in the CF intestine. Databases searched will include PubMed, CINAHL, MEDLINE and the Cochrane library from 1939 until a specified date of last search, alongside clinical trial databases for ongoing studies. Search strategies will include various terminology that relates to the primary mechanistic defects of CF, postulated secondary effects of such defects, and symptoms experienced in patients. A full search strategy is outlined in appendix B. One reviewer will apply an inclusion criterion to obtained abstracts. Following agreement from a second reviewer, full-text articles will be sought, and data will be extracted from relevant articles. Disagreements will be resolved with a third reviewer. The quality of data will be assessed by the GRADE criteria. Data will be used to present a narrative, and where possible, quantitative synthesis.

**Discussion:** This systematic review will discuss our current understanding of the underpinning mechanisms of the persisting abnormalities in gut health and motility within CF, addressing potential intricate relationships that further contribute to disease progression within the intestinal tract. Furthermore, we will identify current gaps in the literature to propose directions for future research. A comprehensive understanding of these aspects in relation to intestinal abnormalities will aid future clinical directions.

## Background

### Description of the condition

Cystic fibrosis (CF) is the most common life-limiting, autosomal recessive genetic disorder, in Europe and North America, with an incidence of about 1 in 2500 births, over ten thousand active patients in the UK alone [1], and a significantly higher incidence in Caucasians [2]. The underlying cellular mechanisms that are disrupted are attributable to mutations in the cystic fibrosis transmembrane conductance regulator (*CFTR*) gene, which encodes a cAMP-regulated ATP-cassette binding protein aptly termed the CFTR protein. CFTR functions to transport chloride (Cl-) and bicarbonate (HCO3-) ions at various epithelial sites throughout the body, whilst also modulating other ionic movements, such as the reabsorption of sodium ions (Na+) via the epithelial sodium channel (ENaC) [3]. Such sites of localisation include the respiratory tract, where CFTR mRNA is expressed in nasal, tracheal, and bronchial epithelial cells [4], and the reproductive tract [5]. As in the airways, the gastrointestinal (GI) tract generally expresses higher levels of CFTR mRNA aside from in the stomach. A decreasing gradient of expression can be observed from the duodenum to the ileum (proximal to distal) inside the small intestine, with expression continuing at a lesser yet moderate level within the colon also. Gradients can too be found within the subsection of the intestinal region in question. In both small/large intestinal environments a crypt-villus gradient is observed in which mRNA expression is shown to decrease [6], although this might not be a true representation of protein levels given the short life cycle of migrating epithelial cells from crypts that could potentially retain and stabilise previously translated protein products. Alongside cholangiocytes at the apical surface of the biliary tree [7], both the pancreas and gallbladder also express CFTR at high levels with the latter in a more uniform fashion, whereas the pancreas displays more expression within the intercalating ducts of the acini [8].

Disruption of CFTR production and function in the airway causes dehydration of the overlying mucosa, which is also speculated to occur at alternate epithelial sites, including the intestines. There is increased viscosity of the mucus layer leading to decreased mucociliary clearance in the airway. In the lung, stasis of respiratory secretions causes bacterial infections in the airway which are recurrent and may become chronic, leading ultimately to bronchiectasis and death from respiratory failure. In recent decades, aggressive antibiotic therapy and the use of mucolytics and airway clearance has delayed the progress of bronchiectasis. This has in turn increased life expectancy for people with CF. A shift in disease manifestations has highlighted the prevalence of the gastrointestinal-related problems associated with CF. Pancreatic insufficiency has been a recognised feature of CF since the first description of the condition [9], arising from the disruption of bicarbonate transport and a failure of pancreatic enzyme secretion. This is present in 86% of people with CF in the US [10]. Cystic fibrosis liver disease (CFLD) has a peak time of onset around puberty and may lead to cirrhosis and portal hypertension [11]. A higher prevalence of GI malignancies is seen in older individuals with CF, compared to the general population [12].

### Gut health and motility in CF

Together with the aforementioned manifestations, abnormalities located at the site of the intestinal tract itself present problems for CF patients, with quality of life affected by daily symptoms which can include abdominal pain, flatulence, bloating, and constipation. CF patients may also experience severe gastrointestinal complications that present antenatally or at birth in the form of meconium ileus. Highlighted by the accumulation of thick, sticky meconium at the terminal ileum and often accompanied by microcolon, it affects around 10-20% of newborns with CF [13]. Later in life, a similar phenomenon may occur in the form of distal intestinal obstruction syndrome (DIOS). DIOS describes small bowel obstruction attributable to the accumulation of partly digested fat at the ileo-caecal valve. This occurs in around 5% of adults and children with CF annually [1]. As both conditions are attributable to mechanical stasis and therefore delayed transit of meconium and chyme respectively through the intestinal lumen, this could well correlate with the occurrence of small intestinal bacterial overgrowth (SIBO), as seen in other GI diseases [14]. CF patients already suffer from malabsorption due to failure of pancreatic enzyme and bicarbonate production. This may be further worsened by bile salt precipitation and enhanced microbial deconjugation of secondary bile acids [15], thereby reducing the effectiveness of vitamin absorption in a lipid-dependent manner.

The CF intestine has received increased attention towards the prevalence of chronic inflammation, more recently demonstrated by non-invasive methods of quantifying calprotectin and M2-PK levels within stool analysis as used for other inflammatory conditions of the intestine [16]. In tandem with the induction of a more pro-inflammatory environment in the CF intestines is the presence of a general microbial dysbiosis, that is the changes in the structure of the microbial community that inhabit this site. There is evidence to suggest a relationship between dysbiosis and inflammation, although the levels of bidirectionality within this relationship requires further elucidation. In early childhood, a rise in the relative abundance of *E. coli* in CF patients significantly correlates with levels of faecal calprotectin [17]. Some species have been assigned as key to optimal gut health, such as prominent members within short-chain fatty acid metabolism and resultant butyrate production, which alongside various other beneficiaries to the host promotes the suppression of localised inflammatory responses by Treg cell expansion [18]. Such species regularly observed in reduced relative abundance in CF includes primary degraders such as *Ruminococcus bromii* and species of *Bifidobacterium*, alongside secondary degraders *Faecalibacterium prausnitzii* and species of *Eubacterium* and *Roseburia* [19]. The proposed role of the gut microbiome is also extended to include its involvement in the facilitation of colorectal cancer development in the CF population [20].

Many of the previous treatments and lifestyle options implemented to control disease-associated complications, such as repeated prophylactic antibiotic administration to control respiratory infection and consuming a diet high in fat and calorific content to increase low body mass index (BMI), are postulated to contribute towards the changes in motility, and health, observed. For example, despite the intervention of pancreatic enzyme replacement therapy (PERT), poor responses to treatment for a multitude of reasons can still render fat absorption incomplete [21]. The administration of antibiotics is often a ‘double-edged sword’, which can result in the emergence of opportunistic pathogens and pathobionts during the course of microbial resilience. CFTR modulator therapies have emerged in recent years with the purpose of correcting the underlying CFTR protein defects within processing and gating at the cell surface. Subsequent to the rescue of CFTR function, preliminary evidence also suggests shifts in gut microbiome composition upon administration alongside other clinical improvements in patients [22].

This systematic review aims to comprehensively address our current understanding of the primary consequences of CFTR dysfunction and their subsequent secondary effects in the context of poor intestinal motility, physiological health, and relating symptoms in the CF population. Further, we will consider intricate relationships that further aggravate the conditions. We will also consider gaps within knowledge and consider the current limitations of study within the various animal models and human sampling methods employed.

### Description of the interventions

We will consider any intervention or study that addresses the prevalence and/or role of the underpinning mechanisms and their secondary persisting effects that lead to negative impacts on gut health and motility in CF. These are outlined in the inclusion criteria.

### Potential impact of this systematic review

As the life expectancy of the CF population continues to rise with improvements in respiratory outcomes of disease, the incidence of intestinal symptoms and manifestations are predicted to increase. Thus, a comprehensive review on the underpinning mechanisms contributing towards these recurrent issues in the CF population is warranted, allowing for potential targets of therapeutic intervention. Most therapies currently in clinical use for the gut symptoms of CF have been repurposed from other disease indications and have not been developed with a knowledge of the mechanisms underpinning gastrointestinal disease in CF.

## Objectives

To conduct a systematic search of the literature to identify the underpinning mechanisms impacting gut health, motility complications, and relating symptoms in CF. This includes primary consequences of CFTR dysfunction and their subsequent secondary effects contributing towards the development of poor intestinal motility, inflammation and other intestinal-based physiological complications.

## Methods

### Criteria for considering studies for this review

#### Types of studies

The review will include all studies investigating the underpinning mechanisms impacting gut health, motility, and relating symptoms in CF. It will include both published and ongoing studies. General reviews, expert opinions, and case reports will be omitted from the subsequent narrative or quantitative synthesis of information but will be used to attain further primary research data where possible.

#### Types of participants

The review will include studies involving participants of any age with a diagnosis of cystic fibrosis defined as either having two CF-causing genetic mutations, or one or no CF-causing mutations and a sweat chloride test greater than 59mmol/L. *CFTR* mutant animal models that reflect human intestinal physiology in CF disease will be included for selection and discussion.

#### Types of symptoms and intestinal complications in health and motility

Symptoms include poor appetite, abdominal pain, flatulence, bloating, and constipation. Motility complications will include DIOS and meconium ileus. Complications in gut health include malabsorption, intestinal inflammation, intestinal lesions and malignancies.

#### Types of mechanisms and secondary effects considered

Mechanisms include primary consequences of CFTR dysfunction, such as reduced fluid production and the failure of pancreatic enzyme and bicarbonate secretion. Subsequent secondary effects considered are increases in luminal fat content, microbial dysbiosis, SIBO, and GI dysmotility (increased transit time). A full outline of the search strategy is found in appendix A. Any report or article that investigates the prevalence or modulation of such mechanisms and/or secondary effects in the context of intestinal health and motility will be included.

#### Types of comparator

For studies assessing the mechanisms underpinning serious motility complications such as DIOS or meconium ileus, this will include both CF and non-CF controls who do not present with obstructive DIOS or meconium ileus. For studies investigating mechanisms of malabsorption, intestinal inflammation, intestinal lesions and malignancies, this will be healthy controls.

#### Types of data to be presented

##### Primary analysis

- Mechanisms and secondary effects underlying DIOS and meconium ileus in CF, or their modulation to alleviate such manifestations.
- Mechanisms and secondary effects underlying malabsorption, intestinal inflammation, intestinal lesions, intestinal malignancies, or their modulation to alleviate such manifestations.

##### Secondary analysis

- Association of symptoms of GI disorder (poor appetite, abdominal pain, flatulence, bloating, and constipation) with motility disorders (DIOS and meconium ileus), or other physiological complications of the CF intestine (malabsorption, intestinal inflammation, intestinal lesions and intestinal malignancies).
- Investigation of the bidirectional relationships between underpinning mechanisms and secondary effects that further accentuates intestinal complications.

### Search methods

Studies will be identified by searching PubMed, CINAHL, MEDLINE and the Cochrane library. The defined search strategies to be used are listed in appendix B. This includes variations in search terms used to increase sensitivity. Searches will be conducted over the period 1/1/1939 – to present, and only articles published in English will be included. In addition, searches will be completed for ongoing clinical trials within the EU clinical trials register, Australia and New Zealand clinical trials register, ISRCTN and Clinicaltrials.gov. Finally, the reference lists of relevant articles and reviews (that may not meet the inclusion criteria) will be searched.

Search results from either databases or clinical trials registers will initially be formatted into excel for de-duplication before first selection, and later downloaded into Mendeley (v.1.19.4) and checked for duplicates followed by a manual check of remaining results.

## Data collection and analysis

### Selection of studies

Titles and abstracts will initially be scanned by one reviewer against the inclusion criteria, and those not relevant will be excluded. After co-agreement from a second reviewer on primary selection, full text articles will be sought for those articles which had met the inclusion criteria based on title or abstract. For articles where either the full text article is not available, or is an abstract from a conference, the abstracts will be reviewed again. If any meaningful results can be gained from this which meet the inclusion criteria these will be included, otherwise they will be excluded. Following second selection, any articles excluded at this stage will have the reason for exclusion recorded.

Where there is disagreement between the two reviewers at any stage of study selection, discrepancies will be discussed until an agreement is reached. If an agreement cannot be reached, we will ask a third reviewer with no attachment or incentives relating to the study.

### Data Extraction

Relevant data from all the included articles after second selection will be placed into a data extraction form created on google sheets to facilitate comparisons between all studies included. We will analyse the underlying mechanistic themes, their secondary effects, and any potential interactions in the context of contributing towards the onset of motility disorders, or the disruption of intestinal physiology in cystic fibrosis to inform a systematic review.

### Quality Assessment

In order to assess the quality of papers obtained and determine the risk of bias, the GRADE criteria will be utilised for each article that passes the second selection process. This will allow for us to assess the strengths and limitations of the current data from the literature in the context of determining the underpinning CFTR-related mechanisms and secondary effects attributable to negative impacts of motility and health in the intestines of the CF population.

## Data Availability

Due to the nature of this manuscript, supporting data is not available. A copy of the PRISMA-P guidelines is attached in the supplementary materials. This has been used to direct the structure of this systematic review protocol.

## Appendix

### A Criteria for considering studies for this review

#### Inclusion

- Diagnosis of CF: diagnosed on genetic or sweat testing at any age (defined as either having two CF-causing genetic mutations, or one or no CF-causing mutations and a sweat chloride test greater than 59mmol/L). Animal based studies will require the disclosure of the defined genotype eliciting a mutation in the *CFTR* gene.
- Participants can be of any age therefore the search will include both children and adults. Studies which include CF patients where intestinal motility and other physiological complications are included as a specific subgroup but analysed individually will also be included.

#### Type of intervention

- Any investigation into the underpinning mechanisms and their secondary effects within CF in the context intestinal motility and other intestinal-based physiological complications. This could be direct prevalence reporting of mechanistic issues and their secondary effects, or further this could be a direct study into the modulation of such mechanisms.
- **Prevalence**: Direct reporting of Pancreatic status, *CFTR-*dependent fluid secretion, SIBO implied by hydrogen/methane breath testing, microbial dysbiosis determined by metagenomic analysis, intestinal inflammation by detection of defined markers, dietary fat consumption, capsule endoscopy scores, or patient questionnaires/reporting of symptoms.
- **Modulation**: Any study or intervention utilising probiotics/prebiotics, antibiotics, restoration of wild-type CFTR function, modulation of defective CFTR, alterations in dietary habits, therapies to increase gut motility, or PERT to increase delivery of enzymes to small intestine.
- Systematic reviews, RCT and observational studies
- Studies 1/1/1939 – to present
- Published and currently ongoing trials

#### Exclusion

- Any study where mechanism(s) and their postulated secondary effects underpinning motility complications (DIOS or meconium ileus), malabsorption, intestinal inflammation, intestinal lesions and intestinal malignancies were not the focal point of study or analysed individually as part of a larger CF study.
- Any study solely comparing *CFTR* genotype to the occurrence of intestinal abnormalities and symptoms in CF.
- Any study looking at the mechanisms underlying inflammation, intestinal mucosal damage (lesions), malabsorption, malignancies, or other physiological changes in the intestines <u>not</u> in the context of CF – e.g. Crohn’s disease or Ulcerative colitis.
- Any study looking at the mechanisms underlying dysmotility not in the context of CF – e.g. Hirschsprung disease or meconium ileus relating to SGA preterm infants.
- Studies involving non-CF pancreatic insufficiency, such as chronic pancreatitis.
- Studies investigating alternate pathways or modifier genes in the aetiology of cystic fibrosis intestinal complications.

### B Search Criteria

Date of search: 1-1-1939 to present

Search terms to be used:

1. Pancreatic
2. Enzyme
3. Pancreas
4. PERT
5. Pancreatic insufficiency
6. Pancreatitis
7. Bicarbonate
8. HCO3
9. Mucus
10. CFTR dysfunction
11. CFTR
12. Restoration
13. Modulator
14. Ivacaftor
15. Lumacaftor
16. Tezacaftor
17. Elexacaftor
18. Orkambi
19. Symkevi
20. Trikafta
21. Kaftrio
22. Symdeko
23. Lubiprostone
24. Dysmotility
25. Motility
26. Transit
27. Small bowel transit
28. Orocecal transit
29. Orocaecal transit
30. Anti-spasmodic
31. Antispasmodic
32. Dysbiosis
33. Microbiome
34. Microbiota
35. Flora
36. Microflora
37. Probiotic
38. Prebiotic
39. Antibiotic
40. Fat
41. Lipid
42. Diet
43. SIBO
44. Small intestinal bacterial overgrowth
45. Small intestine bacterial overgrowth
46. Small bowel bacterial overgrowth
47. Appetite
48. Abdominal
49. Abdomen
50. Flatulence
51. Bloating
52. Constipation
53. Inflammation
54. Inflammatory
55. DIOS
56. Distal intestinal obstruction
57. Distal intestine obstruction
58. Ileocecal
59. Ileocaecal
60. Ileo-cecal
61. Ileo-caecal
62. Meconium ileus
63. Cystic fibrosis
64. CF
65. 1 OR 2 OR 3 4 OR 4 OR 5 OR 6 OR 7 OR 8 OR 9 OR 10 OR 11 OR 12 OR 13 OR 14 OR 15 OR 16 OR 17 OR 18 OR 19 OR 20 OR 21 OR 22 OR 23 OR 24 OR 25 OR 26 OR 27 OR 28 OR 29 OR 30 OR 31 OR 32 OR 33 OR 34 OR 35 OR 36 OR 37 OR 38 OR 39 OR 40 OR 41 OR 42 OR 43 OR 44 OR 45 OR 46
66. 47 OR 48 OR 49 OR 50 OR 51 OR 52 OR 53 OR 54 OR 55 OR 56 OR 57 OR 58 OR 59 OR 60 OR 61 OR 62
67. 63 OR 64
68. 65 AND 66 AND 67

### Databases to be searched

**PubMed:** [Title/Abstract]

**CINAHL:** AB

**MEDLINE:** mp.

**Cochrane:** ti,ab,kw

### Clinical trials registers (EU clinical trials register, Australia and New Zealand clinical trials register, ISRCTN and Clinicaltrials.gov)

1. Appetite
2. Abdominal
3. Abdomen
4. Flatulence
5. Bloating
6. Constipation
7. Inflammation
8. Inflammatory
9. DIOS
10. Distal intestinal obstruction
11. Distal intestine obstruction
12. Ileocecal
13. Ileocaecal
14. Ileo-cecal
15. Ileo-caecal
16. Meconium
17. Cystic fibrosis
18. 1 OR 2 OR 3 4 OR 4 OR 5 OR 6 OR 7 OR 8 OR 9 OR 10 OR 11 OR 12 OR 13 OR 14 OR 15 OR 16 AND 17

